# Association of fibrinogen α_E_, fibrinogen γ′, and sialylated fibrinogen with development of ischemic stroke in patients with recently diagnosed type 2 diabetes

**DOI:** 10.1101/2024.12.19.24319391

**Authors:** Nicoline Daugaard, Else-Marie Bladbjerg, Helene Matilde Lundsgaard Svane, Reimar Wernich Thomsen, Jens Steen Nielsen, Yaseelan Palarasah, Moniek P.M. de Maat, Anna-Marie Bloch Münster

**Author notes:** <u>Corresponding author:</u> Else-Marie Bladbjerg, MSc, PhD, Unit for Thrombosis Research, Section of Clinical Biochemistry, Department of Clinical Diagnostics, University Hospital of Southern Denmark, Esbjerg, Denmark, Institute of Regional Health Research, University of Southern Denmark Finsensgade 35, 6700 Esbjerg, Denmark.

## Abstract

**Background:** Stroke is a major cause of death globally, especially in type 2 diabetes (T2D) patients. Fibrinogen is known to predict stroke risk, but fibrinogen is a highly variable protein and we hypothesized that fibrinogen variants can improve stroke prediction.

**Objectives:** To investigate the association of total fibrinogen and fibrinogen variants with risk of ischemic stroke in T2D patients.

**Methods:** In a nested case-control study with a median follow-up of 4.1 years, we included 144 T2D patients with ischemic stroke (cases) and 144 matched T2D patients without ischemic stroke (controls). We measured total fibrinogen, absolute and relative (ratio between variant and total fibrinogen) levels of three fibrinogen variants (fibrinogen α_E_, fibrinogen γ′, and sialylated fibrinogen) and compared levels between cases and controls. We used logistic regression to determine the association with stroke risk.

**Results:** Total fibrinogen and absolute levels of fibrinogen α_E_, fibrinogen γ′, and sialylated fibrinogen were higher in stroke cases than controls (total fibrinogen 3.55 and 3.44 g/l; fibrinogen α_E_: 8.95 and 8.55 µg/ml; fibrinogen γ′: 638 and 626 µg/ml; sialylated fibrinogen: 1.32 and 1.18 arbitrary units). Absolute levels of fibrinogen positively associated with risk of stroke, both for total fibrinogen (highest vs lowest tertile; adjusted odds ratio (OR) 1.9 (95% CI 0.9-4.2)), fibrinogen γ′ (OR 1.8 (0.8-3.8)), and sialylated fibrinogen (OR 2.5 (1.1-5.8)). Relative levels of fibrinogen variants did not convincingly associate with stroke risk.

**Conclusion:** Patients with T2D who developed stroke had increased levels of total fibrinogen, fibrinogen α_E_, fibrinogen γ′, and sialylated fibrinogen compared with T2D controls. Total fibrinogen and absolute, but not relative, levels of fibrinogen γ′ and sialylated fibrinogen prospectively associated with a 2-fold increased risk of ischemic stroke.

## Introduction

Ischemic stroke is caused by occlusion of a cerebral artery and is globally the second leading cause of death. In Denmark, patients with type 2 diabetes (T2D) have 40% higher risk of developing ischemic stroke than people without T2D ^1^. During the formation of a thrombus, the conversion of fibrinogen to fibrin plays a central role, since the fibrin network stabilizes the thrombus. Fibrinogen has been identified as a risk factor for ischemic stroke ^2^, and since patients with T2D have increased levels of fibrinogen, we hypothesize that this can contribute to their high stroke risk ^3^. Fibrinogen is an acute phase glycoprotein involved in coagulation and inflammation. It is mainly synthesized by hepatocytes and it circulates in plasma at concentrations of 2-4 g/l. It is a 340 kDa protein, consisting of three pairs of chains (2Aα, Bβ and γ) held together by disulfide bridges. In the coagulation cascade, fibrinogen is cleaved by thrombin, and fibrinopeptides are released. This leads to formation of fibrin monomers that form fibers and a fibrin network ^4^.

In a systematic review we recently summarized literature on the association between fibrinogen levels and stroke risk in T2D patients ^5^. The studies did not show consistent associations, which may be partly explained by differences in study participants, designs and low sample sizes. This lack of consistency indicates that we do not yet fully understand the role of fibrinogen in stroke risk of T2D patients. One interesting aspect is the role of fibrinogen variants since their characteristics vary. Variants of the fibrinogen molecule can arise by alternative splicing during gene expression or posttranslational modifications. Both types of fibrinogen variants affect the fibrin clot network, making them highly relevant to explore in relation to stroke in T2D patients ^6^.

One of these fibrinogen splice variants is fibrinogen α_E_ (or fibrinogen-420) with a 236 amino acid extension of the globular C-terminal domain on one or both α-chains (α_E_/α or α_E_/α_E_) ^7^, accounting for 0.5-3% of total fibrinogen in a healthy population ^8,9^. To our knowledge, fibrinogen α_E_ has not previously been measured in patients with T2D or stroke ^9^. Another alternatively spliced fibrinogen variant is fibrinogen γ′ ^10^. Fibrinogen γ′ differs at the C-terminus of the γ-chain, where four carboxy-terminal residues are substituted with a unique sequence of 20 amino acids. Fibrinogen γ′ in the heterodimeric form (γ/γ′) comprises 8-15% of total plasma fibrinogen ^11^ and has been associated with both diabetes mellitus and stroke ^12–14^. Fibrinogen is post-translationally modified by glycosylation, where glycans are enzymatically attached to fibrinogen. Glycosylated fibrinogen is of particular interest in relation to the pathogenesis of T2D ^15^, and fibrinogen has glycosylation sites on each of the three polypeptide chains ^16,17^. When sialic acids are added terminally to the glycans, fibrinogen is sialylated ^15^.

We hypothesized that total fibrinogen and three fibrinogen variants (fibrinogen α_E_, fibrinogen γ′, and sialylated fibrinogen) are associated with the risk of ischemic stroke in recently diagnosed T2D patients. We investigated this in a large prospective cohort study of patients with recently diagnosed T2D followed with respect to stroke development.

## Methods

Because of the sensitive nature of the data collected for this study, requests to access the dataset from qualified researchers trained in human subject confidentiality protocols may be sent to Jens Steen Nielsen.

### Subjects and study design

Danish Centre for Strategic Research in Type 2 Diabetes (DD2) is an ongoing nationwide cohort followed prospectively. Patients with recently diagnosed T2D, clinically diagnosed according to Danish guidelines, are informed about DD2 by their doctor. Interested participants are enrolled at either their general practitioners (estimated 80% of recruited participants) or by hospital outpatient diabetes clinics (estimated 20%) ^18^. The first patient was enrolled in November 2010, and medio 2018 the DD2 cohort consisted of 7936 people with T2D. Baseline blood, urine, and interview data were obtained at the DD2 enrollment visit^19^. Updated baseline and interview data were obtained as part of the International Diabetic Neuropathy Consortium (IDNC) project among DD2 patients who were alive in 2016. The cohort is followed prospectively and linked with different Danish health registries such as Danish Diabetes Database for Adults (DDDA), the Danish National Patient Registry (DNPR), the National Prescription Registry (NPR) and the Danish Civil Registration System (CPR) ^20^.

We present a nested case-control study using data from the DD2 cohort. The study included patients enrolled in DD2 between 2010 and 2017. Patients were excluded from the study if they were diagnosed with T2D, had redeemed a glucose-lowering drug, or had been enrolled in the DDDA database more than four years before DD2 enrollment, had cancer or atrial fibrillation less than one year before enrollment, or were treated with anticoagulants less than 100 days before enrollment as described in the flow chart in Figure 1. On the date of the first stroke (index date), each DD2 stroke case was matched on age (± 1 year), sex, and date of DD2 enrollment (± 1 year) to a diabetes control without a history of stroke. Furthermore, diabetes controls were required to be alive and enrolled in the DD2 database on index date. Following these criteria, we identified a total of 153 first-ever ischemic stroke patients and 153 matching diabetes controls eligible for baseline plasma analysis. It was not possible to analyze plasma in nine of the study subjects due to lack of citrate plasma in the DD2 biobank, leaving 144 complete matched pairs of cases and diabetes controls for analysis. The DD2 patients were followed prospectively for a median of 4.1 years (25-75 percentile 2.7-5.1 years) with respect to development of stroke (International Classification of Diseases (ICD)-10 codes: I60-64, G45.3, G45.8, and G45.9). These ICD codes have a high positive predictive value for stroke diagnoses in DNPR ^21^.

**Figure 1.**
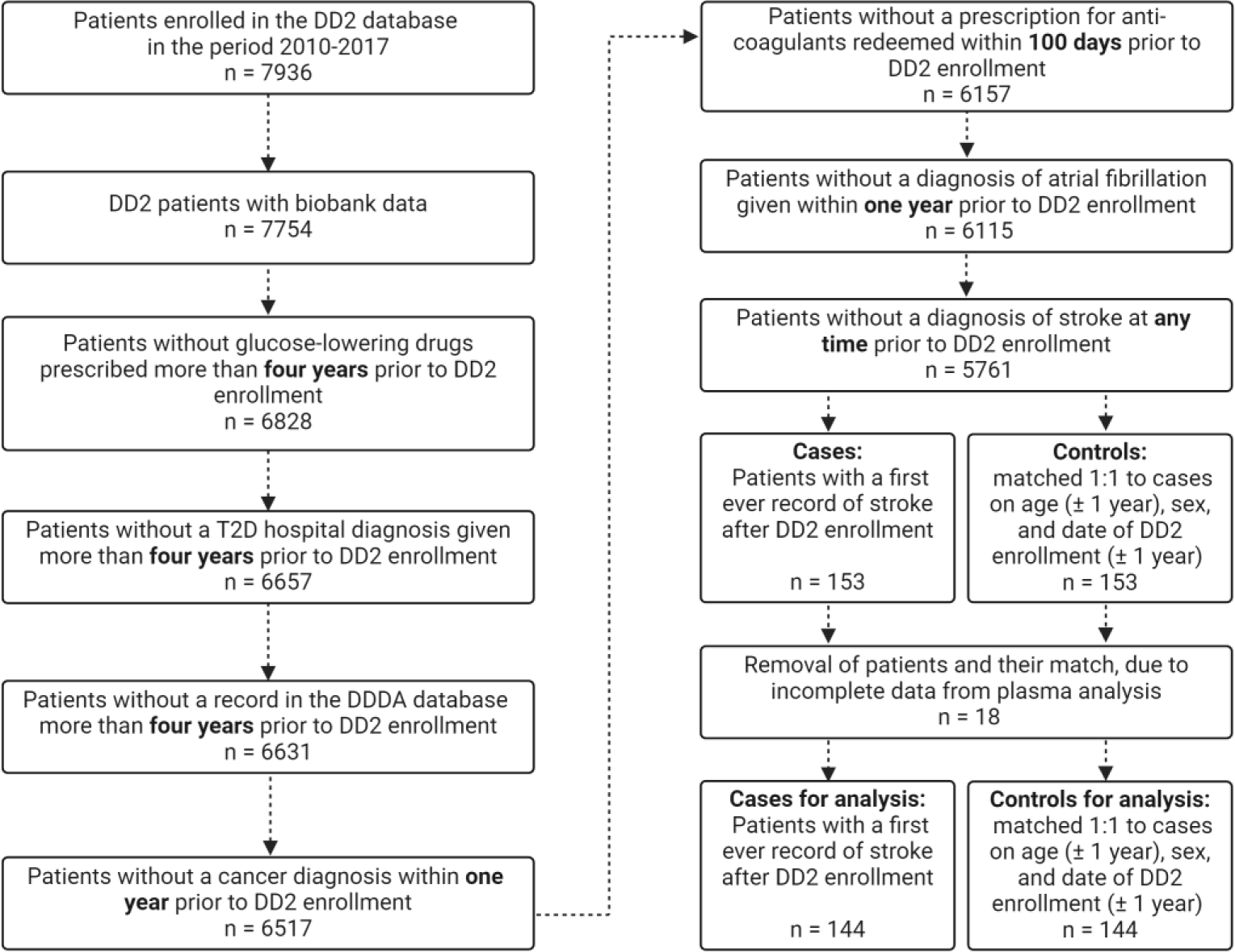
Flowchart of selection of 144 cases with ischemic stroke and 144 matched diabetes controls. Abbreviations: DD2, Danish Centre for Strategic Research in Type 2 Diabetes; DDDA, Danish Diabetes Database for Adults.

To compare levels of fibrinogen variants in diabetes patients and healthy individuals, a group of 120 blood donors was also included in the study. The donors consisted of 60 women and 60 men, who did not use medicine or oral contraceptives. The blood donors were compared to the diabetes controls (n=144) in a case-control design.

### Ethics

The Regional Ethics Committee in the Region of Southern Denmark approved the project (project ID S-20200028), and a data processing agreement (13/11544) was accepted by DD2 and University Hospital of Southern Denmark, Esbjerg. The corresponding author has full access to all the data in the study and takes responsibility for its integrity and the data analysis.

### Blood sampling

#### DD2 participants

Blood sampling took place between 2010 and 2017 at DD2 enrollment. To standardize the blood sampling procedure, a DD2 sampling kit, including manual and video instructions, was sent to the hospital outpatient clinics and general practitioners. Patients arrived in the morning after eight hours of fasting, prior to taking their morning medication. Blood was drawn in vacutainer tubes (2.7 mL trisodium citrate for our analyses) and centrifuged for 10 minutes at 1500-2000 x g at room temperature within two hours after sampling. Plasma was aliquoted and mailed overnight at room temperature to a biobank, where the samples were stored at −80°C until analysis ^19^.

#### Blood donors

Blood from donors was collected in 2.7 mL trisodium citrate vacutainer tubes (Becton Dickinson, Plymouth, UK, ref: 363048) and centrifuged at 2000 x g for 20 minutes. Plasma was aliquoted and stored at −80°C until analysis.

### Fibrinogen analyses

#### Total fibrinogen

Samples were thawed in a water bath at 37°C and case and matching diabetes control were analyzed in the same run. The concentration of total fibrinogen in plasma was determined by nephelometry using the Behring Nephelometer II (Siemens Healthcare Diagnostics Products GmbH, Marburg, Germany) according to instruction of the manufacturer. In brief, complexes are formed with fibrinogen and anti-fibrinogen antibodies, and the intensity of scattered light is proportional to the concentration of fibrinogen. The coefficient of variation (CV) was 3% for within-laboratory variation and for repeatability. The fibrinogen variants described below were expressed both absolute and relative to the concentration of total fibrinogen.

#### Fibrinogen α_E_

Fibrinogen α_E_ was measured in plasma by an in-house enzyme-linked immunosorbent assay (ELISA), with anti-fibrinogen EPR2919 α-chain antibody (Abcam, Amsterdam, Netherlands) as capture antibody and biotinylated polyclonal rabbit fibrinogen antibody (Dako Denmark Aps) as detection antibody. Recombinant human fibrinogen α_E_ (Fibriant, Leiden, the Netherlands) was used as calibrator, and the CV was 17% for within-laboratory variation and 3% for repeatability.

#### Fibrinogen γ′

Plasma levels of fibrinogen γ′ were measured by an in-house ELISA as developed by Pedersen et al. ^22^ using mouse anti-human fibrinogen γ′ monoclonal IgG (19-5-1) as capture antibody and polyclonal rabbit anti-human fibrinogen as detection antibody (Dako Denmark, Glostrup, Denmark). Fibrinogen γ′ (P2 Fib, Enzyme Research Laboratories, South Bend, USA) was used as calibrator. The CV was 7% for within-laboratory variation and 3% for repeatability.

#### Sialylated fibrinogen

For levels of sialylated fibrinogen a new ELISA was developed in this study. A MaxiSorp nunc-immuno plate (ThermoFisher Scientific, Roskilde, Denmark) was coated with 4 µg/ml polyclonal rabbit anti-human fibrinogen antibody (Dako, Glostrup, Denmark Aps) in coating buffer over night at 4°C in a humidity chamber. The following incubation steps were performed at room temperature and under gentle stirring, and the plate was washed four times in PBS-Tween buffer between incubations: The plate was blocked for 30 minutes with dilution buffer (PBS-Tween + 1% bovine serum albumin). Samples, controls, and standard human plasma (Siemens Healthcare Diagnostics Products GmbH, Marburg, Germany) were diluted in dilution buffer and applied in duplicates and incubated for two hours. Next, biotinylated Sambucus Nigra Lectin (SNL) (1:750) (Vector Laboratories, Newark, USA) was added for detection of sialic acids, and the plate was incubated for two hours. Streptavidin horseradish peroxidase (HRP)-conjugate (Invitrogen^TM^, Rockford, USA) diluted 1:3000 in dilution buffer was added and incubated for one hour. The plate was visualized by incubating with 1-Step^TM^ 2,2’-Azinobis [3-ethylbenzothiazoline-6-sulfonic acid]-diammonium salt (ABTS) substrate solution (Thermo Fisher Scientific, Rockford, USA) for 40 minutes. The reaction was stopped by adding 1% sodium dodecyl sulfate solution. The plate was read at 405 nm using a Sunrise absorbance microplate reader (Tecan Group Ltd., Männedorf, Switzerland). Levels of sialylated fibrinogen were calculated by a four parameter marquardt curve fitting (MagellanTM Data Analysis Software v. pro 7.3 for PC, Tecan group Ltd.) of the reference curve made from standard human plasma. Since no standard is available for sialylated fibrinogen, the results were expressed in arbitrary unit (AU), where 1 AU = level in pooled normal plasma. Plasma samples were diluted 1:20.000, and the CV was 13% for within-laboratory variation and 6% for repeatability. Specificity of SNL was tested by incubation over night with α2-3,6,8,9-Neuraminidase, *arthrobacter ureafaciens*, Recombinant, *E. coli*. (Merck KGaA, Darmstadt, Germany) at 37°C. Neuraminidase catalyzes the hydrolysis of terminal sialic acids on glycoproteins, removing the binding sites for SNL and the signal in the ELISA.

### Other measurements

At DD2 enrollment, waist and hip circumferences were measured directly on the skin and waist-hip ratio (WHR) was calculated. Data on alcohol consumption and physical activity levels were collected during an interview using predefined questionnaires. Self-reported tobacco smoking, hemoglobin A1c (HbA1c), and blood pressure were measured and collected by the general practitioner to DDDA. BMI (calculated by dividing body weight with the square of the height) was obtained from DD2 enrollment data when possible, and otherwise from DDDA or IDNC data. Date of first diabetes diagnosis was obtained from a combination of data from DD2, DDDA, and NPR. Blood glucose and C-reactive protein (CRP) were measured by an enzymatic hexokinase method (Glucoquant Glucose/HK, Roche Diagnostics, Mannheim, Germany) and an in-house immunofluorometric assay, respectively ^23,24^. Information about any cardiovascular diseases (CVD) and information about use of medication were from DNPR and NPR, respectively ^20^.

### Statistics

The sample size estimation was based on the best examined fibrinogen variant in the literature, fibrinogen γ′. The sample size was estimated to detect a difference of 100 µg/ml in fibrinogen γ′ between ischemic stroke cases and matched diabetes controls ^13,25,26^ at a significance level of 5%, a power of 80%, and a standard deviation of 300 µg/ml ^22^. Sufficient power was reached when 140 patients were included in each group of this nested case-control study.

Baseline characteristics and fibrinogen measures in stroke cases (n=144) and matched diabetes controls (n=144) are presented as medians (25-75 percentiles) or as numbers (percentages) and were compared with a Mann Whitney test or a Chi^2^ test, respectively. We calculated tertiles for each fibrinogen exposure variable based on the observations in the diabetes control population and used conditional logistic regression to estimate odds ratio (OR) of stroke risk for study subjects with exposure values in the middle and upper tertile, relative to study subject with exposure values in the lower tertile. We calculated crude and adjusted OR. We adjusted for smoking, high blood pressure, CVD (ischemic heart disease, atherosclerosis, congestive heart failure, and venous thromboembolism), WHR, HbA1c, physical activity, and alcohol consumption. These were pre-hoc selected as potential confounders based on a directed acyclic graph (DAG) (see Supplemental Material). We cannot rule out that CVD and WHR might be post-exposure variables in some patients, i.e. factors that could have been affected by elevated fibrinogen levels back in time before we measured the variables. Therefore, we repeated the OR calculations without these variables in the model in a sensitivity analysis (see Supplemental Material). Missing data were addressed by including a subcategory “unknown” in the analyses.

We included the continuous fibrinogen exposures as natural cubic splines with four knots in our regression models and plotted the estimated crude OR and adjusted OR as a function of the exposure, with the fifth percentile as reference. Statistical Analysis System (SAS) 9.4 was used for all analyses in the nested case-control study.

Fibrinogen measured in blood donors (n=120) and the diabetes control group (n=144) are presented as medians (25-75 percentiles) and were compared with a Mann Whitney test in GraphPad Prism version 10.2.3 (GraphPad Software). P < 0.05 was considered statistically significant.

## Results

### Patient characteristics at enrollment

Characteristics of age- and sex-matched cases (n=144) and diabetes controls (n=144) at baseline are presented in Table 1. The two groups were comparable with respect to age at diabetes debut and DD2 enrollment, BMI, alcohol consumption, blood pressure, blood glucose, and medication. Stroke cases had higher median values of CRP and HbA1c and a higher WHR than diabetes controls. More cases than diabetes controls were smokers, physically inactive, had diabetes for a longer period before DD2 enrollment, and had prior CVD comorbidity, although this did not reach statistical significance.

**Table 1.** Enrollment characteristic of T2D patients in cases (with stroke) and controls.

| Characteristics | Cases (n = 144) | Controls (n = 144) | p-value |
| --- | --- | --- | --- |
| Age at T2D debut (years) | 62.5 (55.0;68.0) | 64.0 (56.0;69.0) | 0.84 |
| Age at DD2 enrollment (years) | 66.0 (60.0;73.0) | 66.5 (60.0;73.0) | 0.99 |
| Time from diabetes to DD2 enrollment (months) | 11.0 (3.0;25.5) | 8.0 (2.0-22.0) | 0.12 |
| Time from T2D debut to index date (months) | 49.0 (29.0;64.5) | 46.0 (24.5;65.0) | 0.49 |
| Sex (female) | 63 (43.8) | 63 (43.8) | 1.00 |
| WHR (m) | 1.0 (0.9;1.0) | 1.0 (0.9;1.0) | 0.09 |
| WHR guideline (men/women) |  |  | < 0.001 |
| Low: ≤0.95/≤0.80 m | 16 (11.1) | 18 (12.5) |  |
| Moderate: 0.96-1.00/0.81-0.85 m | 14 (9.7) | 39 (27.1) |  |
| High: >1.00/>0.85 m | 114 (79.2) | 87 (60.4) |  |
| BMI<sup>†,‡</sup> (kg/m<sup>2</sup>) | 30.1 (26.2;33.8) | 29.7 (26.3;33.9) | 0.95 |
| Alcohol intake guideline (women/men) |  |  | 1.00 |
| < 14 units/21 units weekly | 133 (92.4) | 133 (92.4) |  |
| > 14 units/21 units weekly | 11 (7.6) | 11 (7.6) |  |
| Physical activity in the year prior to enrollment |  |  | 0.14 |
| Mainly sedentary | 22 (15.3) | 16 (11.1) |  |
| Light exercise at least 4h/week | 97 (67.4) | 90 (62.5) |  |
| Moderate/heavy at least 4h/week | 25 (17.4) | 38 (26.4) |  |
| Smoking<sup>†,‡</sup> |  |  | 0.33 |
| Smokes daily/occasionally | 25 (22.7) | 15 (15.2) |  |
| Ex-smoker | 41 (37.3) | 37 (37.4) |  |
| Never smoked | 44 (40.0) | 47 (47.5) |  |
| CRP (mg/l) | 2.3 (1.0;4.8) | 1.9 (0.7;3.4) | 0.04 |
| HbA1c<sup>†,‡</sup> (%) | 6.6 (6.2;7.2) | 6.4 (6.0;6.8) | 0.03 |
| HbA1c category |  |  | 0.15 |
| <6.5 % | 49 (42.2) | 56 (53.3) |  |
| 6.5-8 % | 49 (42.2) | 40 (38.1) |  |
| ≥8 % | 18 (15.5) | 9 (8.6) |  |
| Blood glucose<sup>†,‡</sup> (mmol/l) | 7.1 (6.5;8.3) | 7.3 (6.7;8.1) | 0.50 |
| High BP (systolic>140 or diastolic>90 mmHg) |  |  | 0.49 |
| No | 70 (60.9) | 68 (65.4) |  |
| Systolic BP (mmHg) | 132 (125;144) | 130 (124;143) | 0.37 |
| Diastolic BP (mmHg) | 80 (74;85) | 80 (74;85) | 0.10 |
| Antidiabetic drugs | 123 (85.4) | 114 (79.2) | 0.16 |
| Lipid lowering drugs | 106 (73.6) | 105 (72.9) | 0.89 |
| Acetylsalicylic acid | 53 (36.8) | 58 (40.3) | 0.54 |
| NSAIDs | 109 (75.7) | 109 (75.7) | 1.00 |
| Any CVD (IHD, atherosclerosis, HF, or VTE) | 42 (29.2) | 31 (21.5) | 0.14 |

Data are presented as median (25;75 percentiles) or counts (percentages). Abbreviations:
BP, blood pressure; BMI, body mass index; CRP, C-reactive protein; CVD, cardiovascular disease; DD2, Danish Centre for Strategic Research in Type 2 Diabetes; HbA1c, hemoglobin A1c; HF, congestive heart failure; IHD, ischemic heart disease; NSAIDs, nonsteroidal anti-inflammatory drugs; T2D, type 2 diabetes; VTE, venous thromboembolism, WHR, waist-hip ratio.
<sup>†</sup>For cases only available in 65% (blood glucose), 86 % (BMI), 72 % (smoking), 76% (HbA1c).
<sup>‡</sup>For diabetes controls only available in 68 % (blood glucose), 88 % (BMI), 65 % (smoking), 69% (HbA1c)

### Fibrinogen at enrollment

Levels of total fibrinogen and the absolute levels of all three fibrinogen variants (fibrinogen α_E_, fibrinogen γ′, and sialylated fibrinogen) were higher in cases than diabetes controls, although the statistical precision was limited for fibrinogen α_E_ and fibrinogen γ′ (Table 2). The relative levels of fibrinogen α_E_, fibrinogen γ′, and sialylated fibrinogen did not differ between cases and diabetes controls.

**Table 2.**
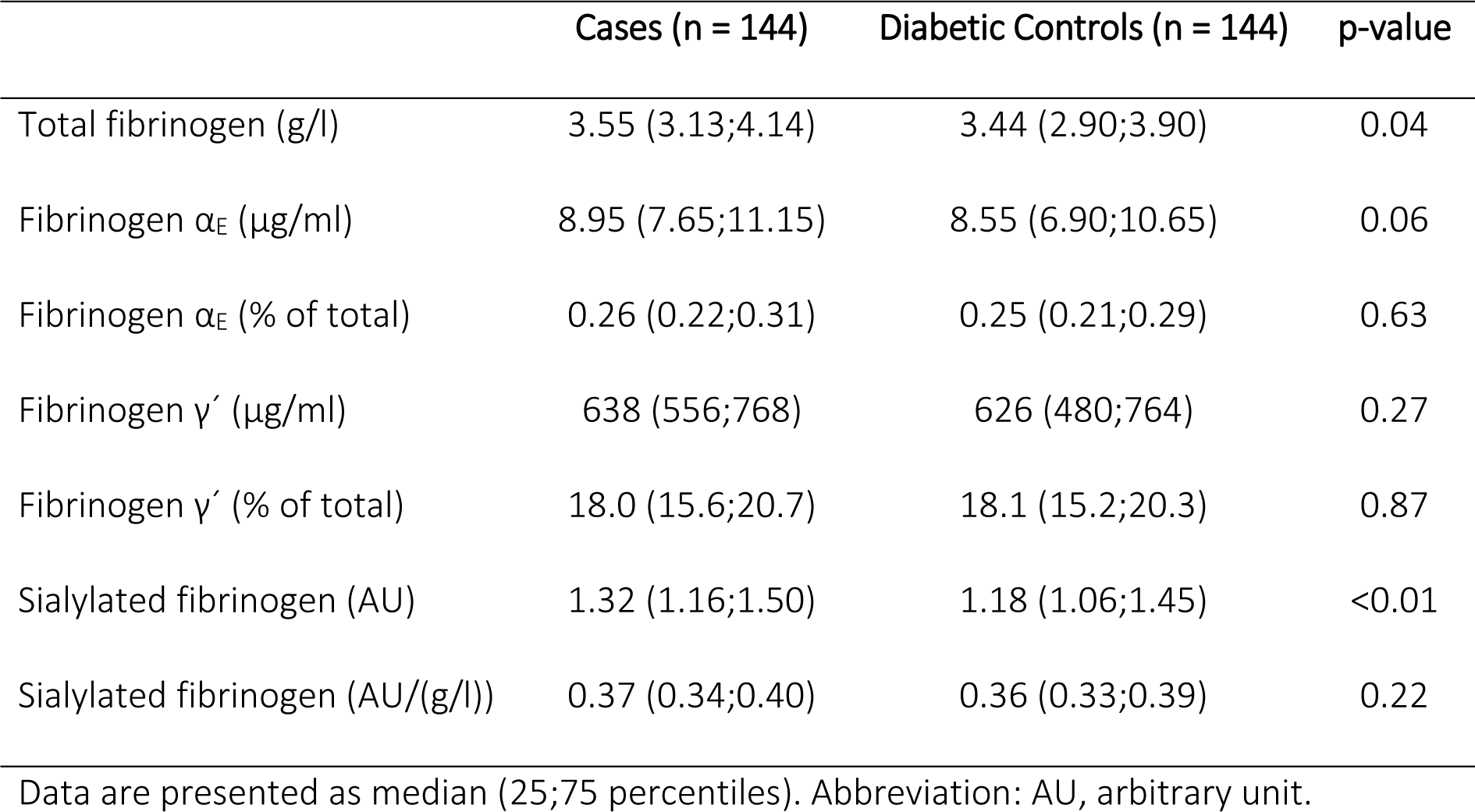
Enrollment measures of fibrinogen in cases (with stroke) and diabetes controls.

### Fibrinogen and stroke risk

Figure 2 presents the OR of ischemic stroke risk for T2D patients based on fibrinogen tertiles. When focusing on unadjusted ORs, patients with total fibrinogen in the highest versus lowest tertile had clearly increased odds of stroke (OR 2.2, 95% CI 1.2-4.3). Absolute levels of fibrinogen γ′ and sialylated fibrinogen were clearly associated with stroke risk in both the highest and middle tertiles, with ORs of around 2 or more compared with the lowest tertile.

**Figure 2.**
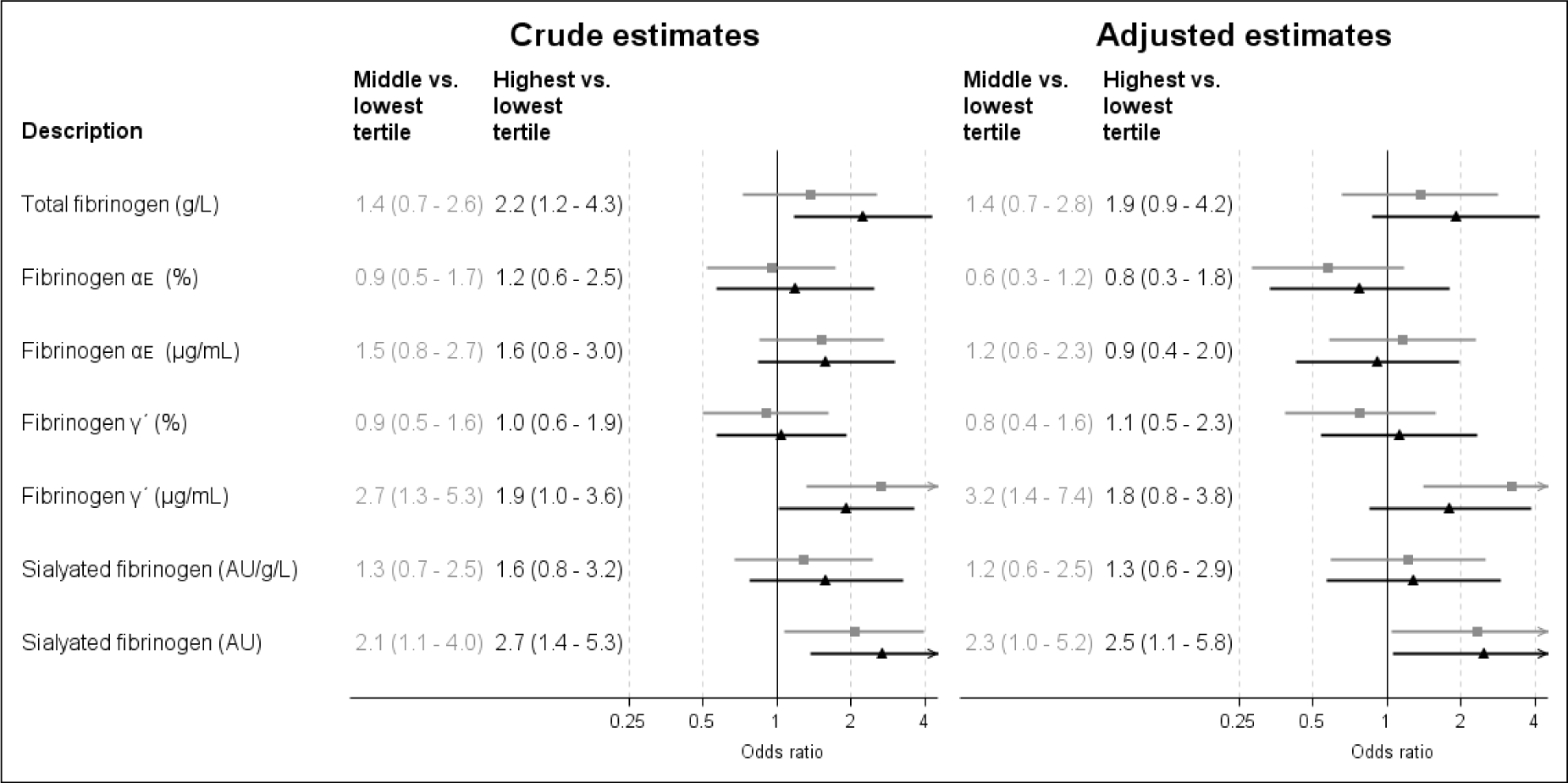
Forest plot of odds ratios for stroke risk for T2D patients based on fibrinogen tertiles. Data is presented with (95% confidence intervals) and as crude and adjusted for smoking, high blood pressure, CVD (ischemic heart disease, atherosclerosis, congestive heart failure and venous thromboembolism), WHR, HbA1c, physical activity, and alcohol consumption. Gray bars represent OR in the second tertile and black bars the third tertile. Abbreviation: AU, arbitrary unit. Lowest tertile: Total fibrinogen: 2.1-3.1 g/l; Fibrinogen α_E_: 4.4-7.5 µg/ml; Fibrinogen α_E_: 0.15-0.23 %; Fibrinogen γ′: 227-529 µg/ml; Fibrinogen γ′: 8.1-16.4 %; Sialylated fibrinogen: 0.74-1.10 AU; Sialylated fibrinogen: 0.25-0.34 AU/(g/l). Middle tertile: Total fibrinogen: 3.1-3.7 g/l; Fibrinogen α_E_: 7.5-9.8 µg/ml; Fibrinogen α_E_: 0.23-0.28 %; Fibrinogen γ′: 529-717 µg/ml; Fibrinogen γ′: 16.4-19.4 %; Sialylated fibrinogen: 1.10-1.35 AU; Sialylated fibrinogen: 0.34-0.38 AU/(g/l). Highest tertile: Total fibrinogen: 3.7-7.2 g/l; Fibrinogen α_E_: 9.8-29.3 µg/ml; Fibrinogen α_E_: 0.28-0.64 %; Fibrinogen γ′: 717-1396 µg/ml; Fibrinogen γ′: 19.4-29.7 %; Sialylated fibrinogen: 1.35-2.94 AU; Sialylated fibrinogen 0.38-0.52 AU/(g/l).

Adjustment for potential confounders did not substantially/importantly reduce the estimated ORs but slightly reduced statistical precision. In adjusted analyses, increasing total fibrinogen was associated with increasing stroke risk (middle tertile OR 1.4 (95% CI 0.7-2.8), highest tertile OR 1.9 (95% CI 0.9-4.2)), although statistical precision was limited. Corresponding stroke ORs for middle and highest tertile of fibrinogen γ′ were 3.2 (1.4-7.4) and 1.8 (0.8-3.8), and for sialylated fibrinogen ORs were 2.3 (1.0-5.2) and 2.5 (1.1-5.8), respectively. Relative levels of fibrinogen variants and absolute levels of fibrinogen α_E_ were not clearly associated with the risk of stroke. Adjusted OR did barely change when suspected post-exposure variables (CVD and WHR) were left out in the sensitivity analysis (see Supplemental Material).

Figure 3 presents plots of adjusted ORs of stroke for continuous levels of total fibrinogen, fibrinogen γ′, and sialylated fibrinogen. These adjusted ORs for stroke risk increased with increasing content of total fibrinogen and sialylated fibrinogen in plasma. The OR for fibrinogen γ′ seemed to have an inverse U-shape with maximum around 600 µg/ml.

**Figure 3.**
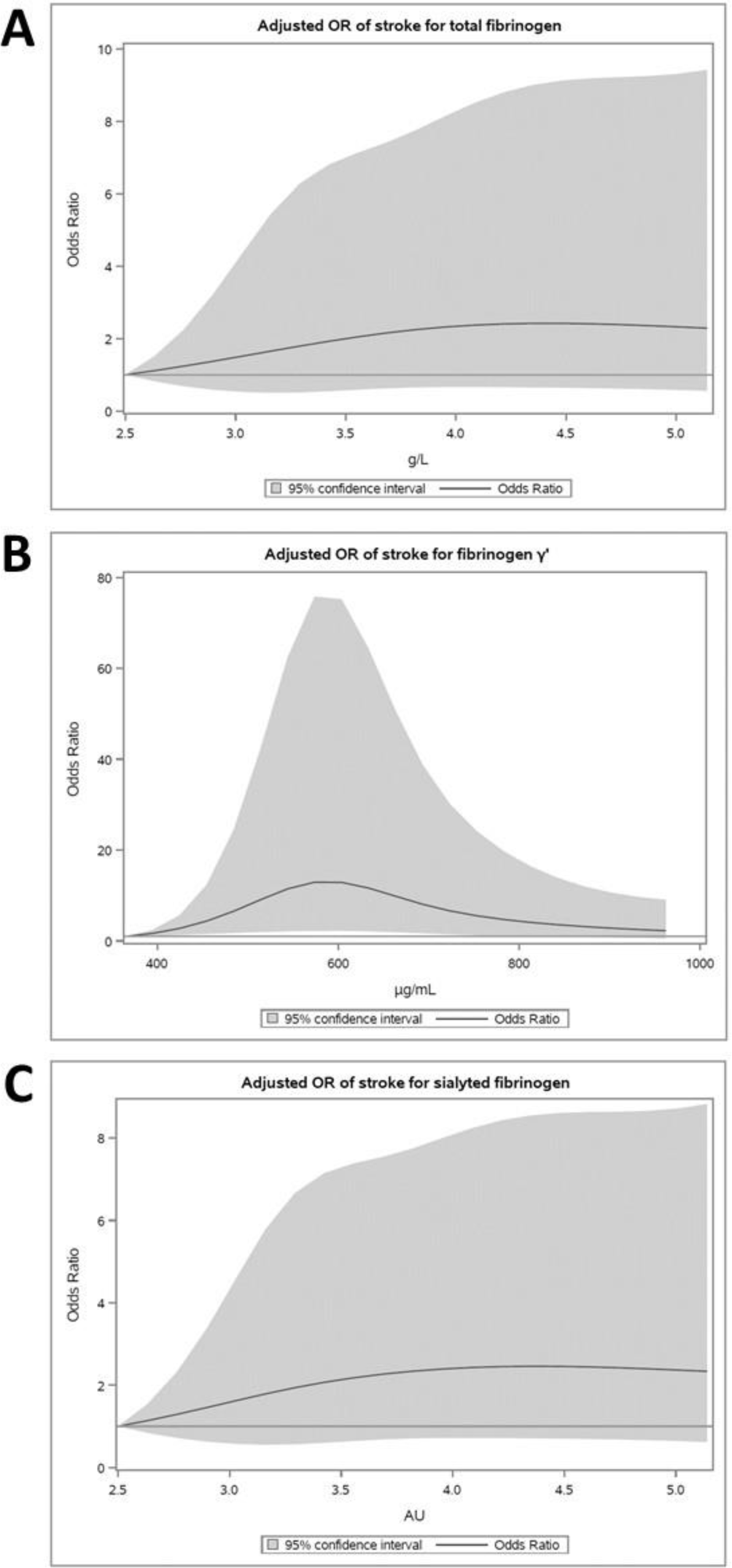
Plots of adjusted odds ratios (ORs) of stroke for (A) total fibrinogen, (B) fibrinogen γ′ and (C) sialylated fibrinogen. Solid grey lines indicate OR estimates and grey shaded areas indicate 95% confidence intervals. ORs are adjusted for smoking, high blood pressure, CVD (ischemic heart disease, atherosclerosis, congestive heart failure and venous thromboembolism), WHR, HbA1c, physical activity, and alcohol consumption. Fibrinogen as continuous variable was modeled with a restricted cubic spline. Abbreviation: AU, arbitrary unit.

### Fibrinogen in blood donors and diabetes controls

The median (25-75 percentiles) age of the blood donors was 27.0 (22.0;35.0) years and body mass index (BMI) was 24.5 (22.3;27.8) kg/m^2^. When comparing levels of fibrinogen variants in blood donors and diabetes controls, we found higher absolute levels and lower relative levels in the diabetes controls (Figure 4).

**Figure 4.**
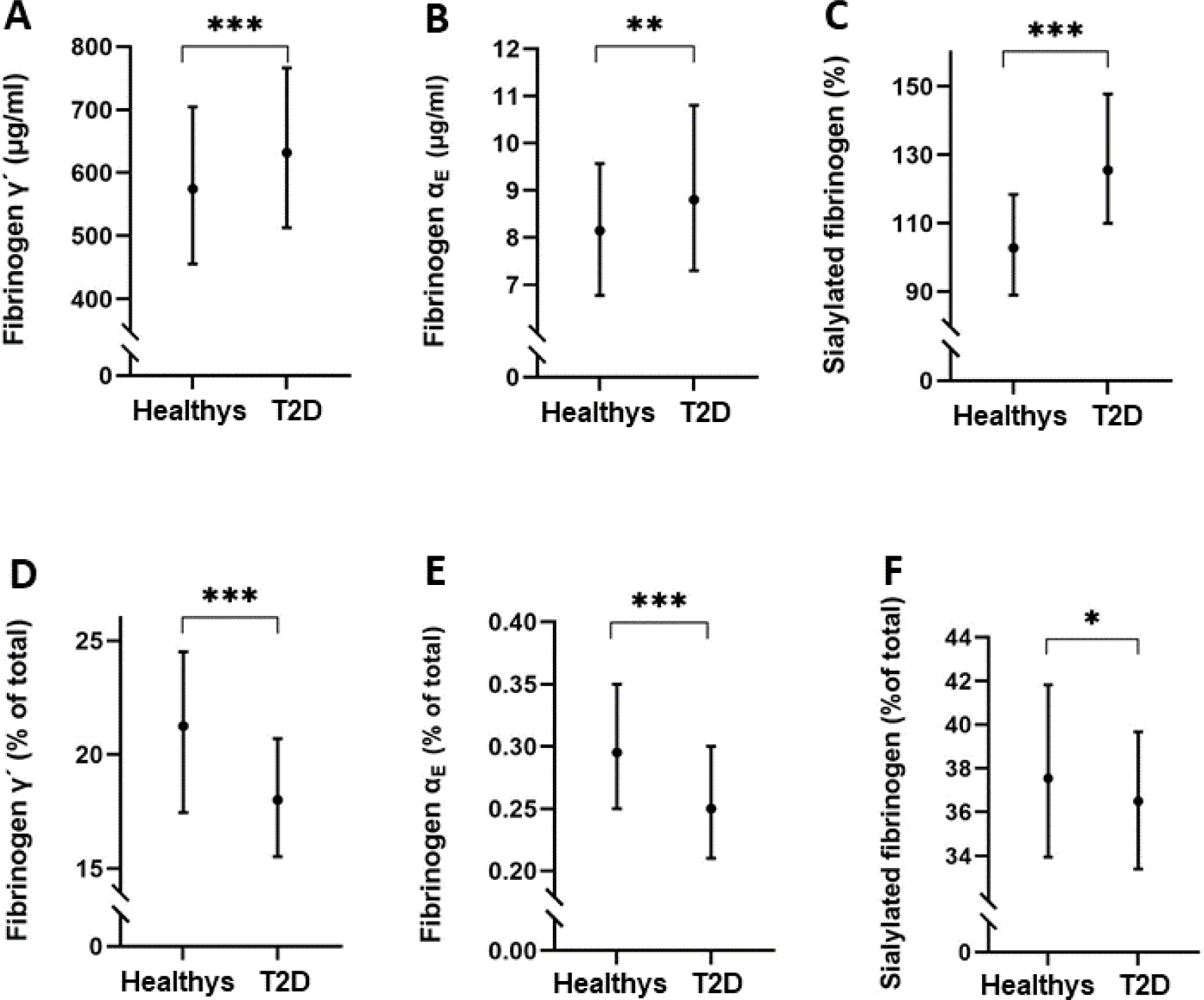
Absolute levels of (A) fibrinogen γ′, (B) fibrinogen α_E_, and (C) sialylated fibrinogen and relative levels of (D) fibrinogen γ′, (E) fibrinogen α_E_, and (F) sialylated fibrinogen in plasma from blood donors (n=120) and diabetes controls (n=144). Data is presented as median with 25-75 percentiles. *p < 0.05, **p < 0.01, ***p < 0.001. Abbreviation: AU, arbitrary unit.

## Discussion

In this nested case-control study, plasma was analyzed from 144 pairs of cases of ischemic stroke and their matched diabetes controls. The study included patients with recently diagnosed T2D, enrolled in DD2 between 2010 and 2017. Concentrations of total fibrinogen and three fibrinogen variants (fibrinogen α_E_, fibrinogen γ′, and sialylated fibrinogen) were compared between cases and diabetes controls, and OR was used to describe their prospective relationship with risk of ischemic stroke. Our main results are that total fibrinogen and absolute levels of fibrinogen α_E_, fibrinogen γ′, and sialylated fibrinogen were higher in stroke cases than diabetes controls at baseline. Furthermore, there was an association between absolute levels of total fibrinogen, fibrinogen γ′, and sialylated fibrinogen at baseline and risk of developing an ischemic stroke in T2D patients. The results confirm our hypothesis that levels of total fibrinogen and fibrinogen variants are associated with stroke risk in T2D patients.

We previously clarified that available literature show inconsistent associations between total fibrinogen levels and risk of stroke in patients with T2D ^5^. Only two studies ^27,28^ have reported an association between levels of total fibrinogen and risk of ischemic stroke in T2D. According to our systematic review ^5^, this study is the second largest study reporting on higher fibrinogen levels in stroke cases than controls in T2D patients ^27,29–31^.

The literature on fibrinogen variants and stroke is very sparse. To our knowledge, no studies have prospectively associated fibrinogen γ′ with an arterial thrombotic event. Two studies reported increased absolute and relative levels of fibrinogen γ′ in patients in the acute phase of ischemic stroke compared to healthy controls ^13,25^. As opposed to this, absolute and relative levels of fibrinogen γ′ decreased in the convalescent phase after ischemic stroke, when compared to the acute phase and to healthy controls ^13^. These studies indicated that fibrinogen γ′ levels are changed at the time of the ischemic stroke, but not how fibrinogen γ′ is affected in the years prior to stroke. In relation to other arterial thromboses, fibrinogen γ′ was increased in patients with myocardial infarction ^32^, coronary artery disease ^33^, and peripheral arterial disease ^34^ in case-control studies. To our knowledge, absolute and relative levels of fibrinogen α_E_ and sialylated fibrinogen were not previously measured in subjects who develop stroke or other arterial events. Stroke was previously observed as an outcome in the DD2 cohort, with increased cumulative incidence with time since enrollment ^35^. We demonstrated that total fibrinogen and absolute levels of three fibrinogen isoforms associate with stroke risk in T2D patients.

Our study did not clarify the mechanisms by which fibrinogen variants associate with stroke risk. The association between fibrinogen γ′ and ischemic stroke can be caused by prothrombotic effects of fibrinogen γ′ on clot formation, as fibrinogen γ′ associates with reduced fibrin fiber diameter, higher density, and increased branching compared with the more common isoform γ/γ fibrinogen ^10^. The association between absolute levels of sialylated fibrinogen and ischemic stroke may be explained by the fact that sialic acids modifies the binding of calcium to fibrinogen. Thereby sialic acids affects fibrin polymerization and clot structure with thinner fibers compared to fibrin without sialic acids suggesting that sialylation makes fibrinogen more prothrombotic ^36^. When we plotted OR against fibrinogen levels as continuous variables, the OR reached a maximum at fibrinogen γ′ levels around 600 µg/ml, although with a large confidence interval. This surprising and interesting observation might suggest that a risk threshold is reached around 600 µg/ml, supported by the decline in OR with increasing fibrinogen γ′ tertiles. As opposed to this, the OR increased from middle to highest tertile of total fibrinogen and sialylated fibrinogen. This indicates a “the higher the worse” relationship between these fibrinogen levels and risk of stroke, until a plateau is reached.

Fibrinogen binds to leukocyte integrin receptors via residues in the C-terminal domain of the γ chain ^37^. The αC-terminal of fibrinogen α_E_ contains an extra leukocyte binding site ^38^, which might increase the pro-inflammatory effect, since the binding between fibrinogen and leukocytes activates the pro-inflammatory NF-κB pathway, leading to transcription of pro-inflammatory cytokines ^39^. Further, since fibrinogen α_E_ lacks a negatively charged polymerization pocket and affects fibrin polymerization with thinner fibers and increased clot stiffness and branching compared with the common α/α fibrinogen ^40^, we expected to see an association between fibrinogen α_E_ and stroke. However, we saw no associations between absolute or relative fibrinogen α_E_ levels and the risk of stroke. It should be noted that the associations between absolute levels of fibrinogen variants and ischemic stroke might only reflect the reported association between total fibrinogen and stroke. In our study, the relative levels of the fibrinogen variants did not relate to stroke, although sialylated fibrinogen showed a tendency. This interesting observation indicates that the more sialic acids on fibrinogen, the higher the risk of stroke in T2D. However, this must be confirmed in a larger study.

Enrollment characteristics of cases and diabetes controls differed with respect to well-known stroke risk factors such as HbA1c, WHR, physical activity, CVD, and smoking ^41–43^. Based on prior knowledge, these risk factors were chosen as confounders in the DAG and adjusted for in the logistic regression together with alcohol consumption and high blood pressure. The adjusted ORs did not change in the sensitivity analysis, where CVD and WHR were left out (see Supplemental Material). This indicates that CVD and WHR should not be considered as post-exposure variables, and they could be included in the adjusted model. CRP also differed among cases and diabetes controls, but its impact on fibrinogen and stroke was adjusted for through the remaining confounders ^44^.

To investigate how healthy people differ from diabetes patients with respect to fibrinogen variants, we compared blood donors with diabetes controls. We found that absolute levels of fibrinogen variants were higher in diabetes controls than in blood donors, whereas relative levels of the fibrinogen variants were higher in blood donors than in diabetes controls. This interesting observation suggests that fibrinogen splice variants and posttranslational modifications are less common in diabetes patients. However, blood donors and diabetes patients differ in many ways, and perhaps older age, higher BMI, or medication use in the diabetes controls, as well as different sample handling procedures, affect the results. More studies are needed in this area. It should be noted that the diabetes controls are not entirely comparable with the general diabetes population, since the diabetes controls can be seen as a more “healthy” and selected part of the DD2 population, who did not develop stroke on index date. Blood donors can likewise be seen as more “healthy” than a standard control population. When comparing blood donors to the total T2D group (n=288) similar results were obtained (results not shown).

As a part of this study, we have developed an ELISA for the quantification of sialylated fibrinogen in plasma. The method is modified from an ELISA for detecting sialylated von Willebrand factor in plasma ^45^. To our knowledge, this is the first time an ELISA has been used to measure sialylated fibrinogen in plasma samples. Until now, determination of sialylated fibrinogen was carried out in two steps. Firstly, fibrinogen had to be isolated from plasma by precipitation with ethanol, amino acids, ammonium sulfate, or by affinity chromatography ^46^. Then the amount of sialic acids was determined on the isolated fibrinogen by colorimetry, fluorometry, enzymatic methods, chromatography, or mass spectrometry ^47^. The strength of the new ELISA is that purification of fibrinogen is not needed, and the method allows for measuring the concentration of sialylated fibrinogen directly in plasma. The assay can be applied in future studies where sialylated fibrinogen is expected to play a role, e.g., in newborns ^48^ and in patients with liver disease ^49^.

One of the methodological strengths of this study is that matched cases and diabetes controls were followed prospectively in Danish registries allowing full follow-up of all participants. Also, participants were recently diagnosed with T2D (<4 years before study inclusion), leaving the diabetic changes in the body at a minimum. Another strength is that fibrinogen was measured in blood samples obtained relatively close to the stroke incidence, reducing the time for impact of other factors. A limitation is that plasma from DD2 patients was mailed overnight at room temperature to the biobank for logistical reasons in this nationwide study ^19^. According to the Clinical and Laboratory Standards Institute (CSLI), plasma stored for more than 4 hours should be frozen ^50^. Since no studies have investigated the stability of fibrinogen in plasma stored overnight at room temperature, we performed a pilot study (n=10) and found no changes in levels of total fibrinogen, sialylated fibrinogen, and fibrinogen γ′, whereas levels of fibrinogen α_E_ were significantly reduced (median reduction of 16%) (results not shown). This indicates that lower levels of fibrinogen α_E_ are reported in this study than if the samples were handled according to CLSI. Even though samples from cases and controls were handled identically, the lack of association between fibrinogen α_E_ and stroke risk must be confirmed in a future study.

In conclusion, total fibrinogen and absolute levels of three fibrinogen variants (fibrinogen α_E_, fibrinogen γ′, and sialylated fibrinogen) were higher in patients with T2D who over the next years developed stroke compared with matched diabetes controls, who did not develop stroke. Furthermore, total fibrinogen and absolute levels of fibrinogen γ′ and sialylated fibrinogen were prospectively associated with risk of ischemic stroke, but fibrinogen αE was not. Patients with T2D had higher absolute levels, but lower relative levels, of the three fibrinogen variants compared to blood donors.

## Data Availability

Requests to access the dataset from qualified researchers trained in human subject confidentiality protocols may be sent to the corresponding author.

## Acknowledgements

We would like to thank Miranda Weggeman from Fibriant BV, Leiden, the Netherlands for valuable advice during the setup of the fibrinogen α_E_ ELISA.

## Sources of funding

This work was supported by a grant from Steno Diabetes Center Odense, funded by the Novo Nordisk Foundation. The Region of Southern Denmark and Karola Jørgensens research foundation also supported the project.

## Disclosures

None.

## Supplemental Material

Figure S1

Table S1

## Non-standard Abbreviations and Acronyms

AU: Arbitrary unit
CSLI: Clinical and Laboratory Standards Institute
DD2: Danish Centre for Strategic Research in Type 2 Diabetes
CPR: Danish Civil Registration System
DDDA: Danish Diabetes Database for Adults
DNPR: Danish National Patient Registry
ICD: International Classification of Diseases
IDNC: International Diabetic Neuropathy Consortium
NPR: National Prescription Registry
SNL: Sambucus Nigra Lectin
SAS: Statistical Analysis System
T2D: Type 2 diabetes

## Notes

### Competing Interest Statement

The authors have declared no competing interest.

### Clinical Trial

The study was not registered, since it is a purely observational study.

### Author Declarations

The Regional Ethics Committee in the Region of Southern Denmark approved the project (project ID S-20200028), and a data processing agreement (13/11544) was accepted by DD2 and University Hospital of Southern Denmark, Esbjerg.

